# Changes in body composition and adiposity following an eight-week health-promotion intervention among adolescents and young adults with type 1 diabetes in Wakiso, Uganda: A controlled before-and-after study

**DOI:** 10.64898/2026.10.03.26364632

**Authors:** Jonathan Nsamba, Safaa Garelnabi, Charles Mwanje, Sheena Arayo, Ronald Tibiita, Karen Helmy Ademun, Ruth Biyinzika

## Abstract

**Background:** Nutrition education and physical activity support are important components of type 1 diabetes mellitus (T1DM) care, but their association with body composition outcomes in routine Ugandan diabetes services remains uncertain. We evaluated a culturally tailored health promotion programme among adolescents and young adults living with T1DM.

**Methods:** We conducted a trial study in Wakiso Health Centre IV, Uganda. Sixty participants aged 13–26 years were allocated through an alternating clinic-week schedule to an intervention group or usual-care control group, with 30 participants per group. The eight-week Active Hearts, Healthy Lives programme combined nutrition education, personalised dietetic counselling, physical activity support, and practical self-management activities. Anthropometric and bioelectrical impedance measurements were obtained at baseline and six months after baseline. Separate linear regression models compared follow-up outcomes, adjusting for the corresponding baseline measurement, age, and sex.

**Results:** Participants had a mean age of 19.45 years (SD 3.02), and 33 (55.0%) were female. Compared with controls, the intervention group had lower adjusted fat mass (mean difference −1.70 kg; 95% CI −2.48 to −0.92) and BMI (−0.72 kg/m²; −1.39 to −0.06), and higher fat-free mass (4.39 kg; 3.93 to 4.84) and muscle mass (1.73 kg; 0.84 to 2.62). These differences reflected modest changes among intervention participants alongside increased fat mass and decreased fat-free mass and muscle mass among controls.

**Conclusion:** Participation in the programme was associated with differences in estimated body composition at six months. Larger randomised evaluations incorporating clinical and functional outcomes are needed to establish whether the programme provides meaningful health benefits.

## Introduction

Care for adolescents and young adults living with type 1 diabetes mellitus (T1DM) requires sustained attention to nutrition, physical activity, and self-management alongside insulin therapy (1). International guidance recognises nutritional support and exercise education as integral components of diabetes care, with advice tailored to developmental needs, family circumstances, and cultural practices (2,3). These considerations are particularly relevant when designing interventions intended to support healthy growth and body composition while remaining practical within participants’ everyday lives.

Body composition provides an important perspective on nutritional status beyond body weight alone (4). Weight and body mass index (BMI) do not distinguish fat mass from fat-free tissue, potentially obscuring differences in the composition of body weight. Earlier research in Uganda by Nsamba and colleagues, among children living with T1DM, identified differences in body composition between children and adolescents with T1DM and matched participants without diabetes (5). In particular, participants with T1DM had a higher fat mass index. These findings provided a rationale for investigating approaches to address adiposity and for measuring fat and fat-free compartments alongside conventional anthropometric outcomes.

Ugandan evidence also highlights nutritional and practical challenges relevant to health promotion among young people with T1DM. Ndahura and colleagues found that 93.2% of caregivers attending six diabetes clinics had low diabetes-related nutrition knowledge, with carbohydrate counting the weakest knowledge domain (6). Their findings supported the need for tailored caregiver nutrition education (4). Among young adults aged 18–25 years in rural Uganda, Sseguya and colleagues documented reduced meal frequency, blood glucose monitoring, and insulin injection frequency, alongside reduced access to food and diabetes support (7). Evidence from other settings demonstrates the importance of addressing the circumstances in which young people manage diabetes. In Kenya, Palmer and colleagues described difficulties adapting dietary practices and daily routines among children with T1DM (8). Together, these findings provide a rationale for combining culturally appropriate nutrition education and physical activity support with practical self-management skills.

Intervention research provides evidence that behavioural support can improve some aspects of diabetes management, although improvements may differ across outcomes. An 18-month randomised trial among young people with T1DM in the United States found that a family-based behavioural nutrition intervention improved dietary quality without improving glycaemic control (9). Exercise guidance similarly emphasises individualised support, including education on glucose monitoring, nutrition, and insulin management around activity (3).

However, these studies do not establish whether a culturally tailored programme delivered within routine Ugandan diabetes care is associated with changes in body composition. Examining fat mass, fat-free mass, and muscle mass alongside weight and BMI can characterise changes that conventional anthropometric measures alone may obscure. We therefore evaluated Active Hearts, Healthy Lives, an eight-week programme combining nutrition education, personalised dietetic counselling, physical activity support, and practical diabetes self-management activities at Wakiso Health Centre IV, Uganda. We compared changes in these outcomes over six months from baseline between adolescents and young adults receiving the programme and those receiving usual diabetes care. This evaluation aimed to provide preliminary evidence to inform locally appropriate health promotion within diabetes services.

## Methods

### Study design and setting

This was a clinical trial to evaluate changes in anthropometric and body composition measures following a culturally tailored health promotion intervention among adolescents and young adults living with type 1 diabetes mellitus. The study took place at Wakiso Health Centre IV, a government-owned health facility in Wakiso District, Uganda, providing diabetes care through the Changing Diabetes in Children programme (10). Participants received either an eight-week health promotion intervention or usual diabetes care. Assessments were conducted at baseline and six months after baseline.

### Participants and allocation

The quantitative analysis included 60 participants receiving diabetes care at the study facility, with 30 participants in each study group. Participants were aged 13–26 years at baseline. The original protocol targeted children and adolescents aged 12 to less than 18 years with diagnosed type 1 diabetes mellitus who were enrolled in the Changing Diabetes in Children programme.

Participants were recruited through the diabetes clinic. Allocation followed an alternating clinic-week schedule. Participants attending designated control sessions entered the control group, whereas those attending intervention sessions entered the intervention group. Group membership was maintained throughout the study, with separate clinic sessions intended to minimise contamination. Allocation was therefore based on the clinic attendance schedule.

### Intervention and usual care

The intervention comprised the eight-week Active Hearts, Healthy Lives programme, designed to support diabetes self-management through nutrition education, physical activity, and practical skills development. Programme development was informed by Social Cognitive Theory (11) and the Health Belief Model (12). Its content was adapted to locally available foods, resources, and cultural preferences.

The programme included education on diabetes management, personalised dietetic counselling, practical demonstrations of glucose monitoring and healthy meal preparation, and structured physical activity sessions. Nutrition activities emphasised practical food choices and meal preparation using locally available foods. Physical activity activities were adapted to participants’ circumstances and available resources. The intervention was delivered by well-trained diabetes nurses and a nutritionist. Participants in the control group continued to receive usual diabetes care at the facility.

### Data collection and outcome measures

Data were collected at baseline and six months after baseline using a structured study questionnaire and physical measurements. Baseline characteristics included age, sex, ethnicity, religion, dietary practices, self-reported physical activity, and clinical measurements. Dietary questions covered daily meal frequency and the addition of extra salt to food. The study questionnaire included a 24-hour dietary recall and a checklist of ten food groups. Each food group was coded according to whether it had been consumed during the preceding day and night. The resulting food-group count ranged from 0 to 10 and represented the number of listed groups consumed.

The outcomes examined in this manuscript were body weight, body mass index, fat mass, fat-free mass, and muscle mass. Participants were measured barefoot and wearing light clothing. Body weight was measured to the nearest 0.1 kg using a Seca portable electronic scale (Model 874, Hamburg, Germany). Standing height was measured to the nearest 0.1 cm using a wall-mounted stadiometer, with participants standing erect.Body composition was assessed using a Tanita DC-430MA bioelectrical impedance analyser (TANITA Corporation, Tokyo, Japan), with participants standing barefoot on the electrode platform. Fat mass, fat-free mass, and muscle mass were recorded in kilograms. Body mass index was expressed in kilograms per square metre. For each outcome, within-participant change was calculated by subtracting the baseline value from the six-month value. Positive values indicated an increase, and negative values indicated a decrease during follow-up.

### Statistical analysis

We conducted analyses using IBM SPSS Statistics, version 31. Continuous variables were summarised as means and standard deviations, and categorical variables as frequencies and percentages. Baseline characteristics were described overall and by study group. For each outcome, baseline values, six-month values, and within-participant changes were summarised separately for the intervention and control groups.

We examined between-group differences at six months using separate multiple linear regression models for body weight, body mass index, fat mass, fat-free mass, and muscle mass. Each model included the six-month outcome as the dependent variable and study group as the explanatory variable of interest, adjusting for the baseline value of the corresponding outcome, age, and sex.

The control group served as the reference category. Study-group coefficients were reported as adjusted mean differences between the intervention and control groups, with 95% confidence intervals and two-sided p-values. Positive coefficients indicated higher adjusted six-month values in the intervention group, whereas negative coefficients indicated lower values.

### Ethical considerations

Ethical approval was obtained from the St. Francis Hospital Nsambya Research Ethics Committee (SFHN-2024-133). Before enrolment, participants received information about the study’s purpose, procedures, potential risks and benefits, and their rights as research participants. Participation was voluntary, and participants could decline participation or withdraw at any time without affecting their access to diabetes care or other services at the facility.

For participants younger than 18 years, written informed consent was obtained from a parent or legal guardian, together with assent from the participant. For participants aged 18 years or older, written informed consent was obtained directly from the participant. Participants and their parents or guardians had opportunities to ask questions before deciding whether to participate.

Privacy and confidentiality were maintained throughout the study. Data collection took place in a private setting, and study identification codes replaced names on questionnaires and measurement records. Consent forms and any records linking identification codes to participants were stored separately from the research data. Paper records were kept in locked storage, while electronic records were password-protected and accessible only to authorised members of the research team.

## Results

### Participant demographics and baseline characteristics

The study included 60 participants, with 30 in the intervention group and 30 in the control group. The mean age was 19.45 years (SD 3.02; range 13–26), and 33 participants (55.0%) were female. More than half were aged 20–26 years (53.3%), while 28.3% were aged 13–17 years and 18.3% were aged 18–19 years. Most participants identified as Baganda (65.0%). Catholic affiliation was the most frequently reported religion (33.3%), followed by Born Again (30.0%), Protestant (20.0%) and Muslim (16.7%).

At baseline, participants in the intervention group were taller and had a higher mean body weight than those in the control group, although their mean BMI was lower. Mean fat mass was 10.88 kg (SD 8.47) in the intervention group and 16.07 kg (SD 7.67) in the control group. Conversely, mean fat-free mass was higher in the intervention group, at 43.15 kg (SD 7.04), compared with 39.48 kg (SD 11.62) in the control group. Mean muscle mass was similar between groups, at approximately 41 kg.

Most participants reported consuming three meals daily (90.0%), and 18.3% reported adding extra salt to food. The mean recorded dietary diversity score was higher in the intervention group than in the control group, at 5.20 (SD 0.92) and 4.37 (SD 1.35), respectively. Physical-activity participation was reported by 56.7% overall, including 63.3% of the intervention group and 50.0% of the control group. Mean baseline systolic and diastolic blood pressures were 122.07 mmHg (SD 14.02) and 74.35 mmHg (SD 9.72), respectively. Participant characteristics are summarised in **Table 1**.

**Table 1.** Participant demographics and baseline characteristics by study group.

| Characteristic | Control (n = 30) | Intervention (n = 30) | Overall (N = 60) |
| --- | --- | --- | --- |
| Age, years | 19.80 (3.39) | 19.10 (2.62) | 19.45 (3.02) |
| 13–17 years, n (%) | 9 (30.0) | 8 (26.7) | 17 (28.3) |
| 18–19 years, n (%) | 4 (13.3) | 7 (23.3) | 11 (18.3) |
| 20–26 years, n (%) | 17 (56.7) | 15 (50.0) | 32 (53.3) |
| Sex, n (%) |  |  |  |
| Female | 18 (60.0) | 15 (50.0) | 33 (55.0) |
| Male | 12 (40.0) | 15 (50.0) | 27 (45.0) |
| Ethnicity, n (%) |  |  |  |
| Baganda | 21 (70.0) | 18 (60.0) | 39 (65.0) |
| Other ethnicities | 9 (30.0) | 12 (40.0) | 21 (35.0) |
| Religion, n (%) |  |  |  |
| Catholic | 9 (30.0) | 11 (36.7) | 20 (33.3) |
| Born Again | 8 (26.7) | 10 (33.3) | 18 (30.0) |
| Protestant | 8 (26.7) | 4 (13.3) | 12 (20.0) |
| Muslim | 5 (16.7) | 5 (16.7) | 10 (16.7) |
| Height, cm | 154.27 (7.18) | 160.77 (8.00) | 157.52 (8.22) |
| Weight, kg | 52.48 (5.97) | 54.03 (9.93) | 53.26 (8.16) |
| BMI, kg/m<sup>2</sup> | 22.11 (2.59) | 21.02 (4.45) | 21.56 (3.65) |
| Fat mass, kg | 16.07 (7.67) | 10.88 (8.47) | 13.48 (8.43) |
| Fat-free mass, kg | 39.48 (11.62) | 43.15 (7.04) | 41.32 (9.70) |
| Muscle mass, kg | 40.99 (7.30) | 40.94 (6.70) | 40.97 (6.94) |
| Meals per day, n (%) |  |  |  |
| Two | 3 (10.0) | 1 (3.3) | 4 (6.7) |
| Three | 27 (90.0) | 27 (90.0) | 54 (90.0) |
| Four | 0 (0.0) | 2 (6.7) | 2 (3.3) |
| Adds extra salt to food: yes, n (%) | 3 (10.0) | 8 (26.7) | 11 (18.3) |
| Dietary diversity score | 4.37 (1.35) | 5.20 (0.92) | 4.78 (1.22) |
| Reports physical-activity participation: yes, n (%) | 15 (50.0) | 19 (63.3) | 34 (56.7) |
| Systolic blood pressure, mmHg | 122.80 (13.71) | 121.33 (14.52) | 122.07 (14.02) |
| Diastolic blood pressure, mmHg | 73.50 (10.42) | 75.20 (9.06) | 74.35 (9.72) |
| Pulse rate, beats/min | 81.33 (8.94) | 80.90 (10.49) | 81.12 (9.67) |

*Values are mean (SD), unless otherwise indicated. BMI, body mass index; SD, standard deviation.*

### Changes in body composition over six months

Mean changes in fat mass, fat-free mass and muscle mass differ between the intervention and control groups **(Table 2).** Mean fat mass decreased by 0.14 kg (SD 2.19) in the intervention group, from 10.88 to 10.74 kg, whereas it increased by 0.72 kg (SD 1.64) in the control group, from 16.07 to 16.79 kg. Recorded fat-free mass increased by 0.98 kg (SD 0.72) in the intervention group and decreased by 3.11 kg (SD 1.40) in the control group. Similarly, mean muscle mass increased by 0.25 kg (SD 1.43) in the intervention group, compared with a decrease of 1.46 kg (SD 2.56) in the control group. Mean body weight decreased by 0.88 kg (SD 1.85) in the intervention group and increased by 0.47 kg (SD 3.93) in the control group. Corresponding changes in BMI were −0.32 kg/m² (SD 0.69) and +0.32 kg/m² (SD 1.68), respectively. Individual weight changes varied more widely in the control group, ranging from −13.10 to +12.70 kg, compared with −7.80 to +3.00 kg in the intervention group.

**Table 2.** Body-composition measurements at baseline and six months, by study group.

| Outcome | Study group | Baseline, mean (SD) | Six months, mean (SD) | Change, mean (SD) |
| --- | --- | --- | --- | --- |
| Fat mass, kg | Control | 16.07 (7.67) | 16.79 (6.53) | +0.72 (1.64) |
|  | Intervention | 10.88 (8.47) | 10.74 (7.30) | -0.14 (2.19) |
| Fat-free mass, kg | Control | 39.48 (11.62) | 36.37 (10.27) | -3.11 (1.40) |
|  | Intervention | 43.15 (7.04) | 44.13 (7.27) | +0.98 (0.72) |
| Muscle mass, kg | Control | 40.99 (7.30) | 39.53 (4.88) | -1.46 (2.56) |
|  | Intervention | 40.94 (6.70) | 41.19 (6.97) | +0.25 (1.43) |
| Weight, kg | Control | 52.48 (5.97) | 52.95 (6.06) | +0.47 (3.93) |
|  | Intervention | 54.03 (9.93) | 53.15 (9.67) | -0.88 (1.85) |
| BMI, kg/m<sup>2</sup> | Control | 22.11 (2.59) | 22.43 (2.34) | +0.32 (1.68) |
|  | Intervention | 21.02 (4.45) | 20.70 (4.41) | -0.32 (0.69) |

*Change was calculated for each participant as the six-month measurement minus the baseline measurement; positive values indicate increases and negative values indicate decreases. Abbreviations: BMI, body mass index; SD, standard deviation.*

### Exploratory adjusted between-group comparisons

Separate linear regression models compared six-month outcomes between groups, adjusting for the corresponding baseline measurement, age and sex. After adjustment for baseline age and sex, six-month fat mass was lower in the intervention group than in the control group, with an adjusted mean difference of −1.70 kg (95% CI −2.48 to −0.92). Conversely, adjusted fat-free mass and muscle mass were higher in the intervention group, with differences of 4.39 kg (95% CI 3.93 to 4.84) and 1.73 kg (95% CI 0.84 to 2.62), respectively **(Table 3).** The adjusted between-group difference in BMI was −0.72 kg/m² (95% CI −1.39 to −0.06). For body weight, the estimated difference was −1.13 kg (95% CI −2.74 to 0.47).

**Table 3.** Exploratory adjusted between-group differences in body composition at six months.

| **Outcome** | **Adjusted mean difference** | **95% confidence interval** | **p-value** |
| --- | --- | --- | --- |
| Fat mass, kg | -1.70 | -2.48 to -0.92 | <0.001 |
| Fat-free mass, kg | +4.39 | 3.93 to 4.84 | <0.001 |
| Muscle mass, kg | +1.73 | 0.84 to 2.62 | <0.001 |
| Weight, kg | -1.13 | -2.74 to 0.47 | 0.162 |
| BMI, kg/m<sup>2</sup> | -0.72 | -1.39 to -0.06 | 0.034 |

*P-values estimated using separate linear regression models adjusted for the corresponding baseline age and sex.*

## Discussion

In this single-centre study of adolescents and young adults living with T1DM in Uganda, participation in an eight-week culturally tailored health promotion programme was associated with lower fat mass and BMI and higher fat-free mass and muscle mass at six months, after adjustment for the corresponding baseline measurement, age, and sex. The body composition differences reflected modest changes among intervention participants alongside increases in fat mass and declines in fat-free mass and muscle mass among controls.

Previous intervention studies provide support for these findings, although results have varied. In a small randomised study of adolescent girls with T1DM, Heyman and colleagues reported increased fat mass in the non-training group and increased fat-free mass in participants undertaking combined aerobic and strength training (13). By contrast, D’hooge and colleagues found improvements in physical fitness and reduced insulin requirements after combined exercise training without significant changes in anthropometric measures (14). More recently, the 24-week Diactive-1 randomised trial reported a between-group difference in lean mass gain of 0.88 kg following an intervention involving progressive resistance training (15). These studies indicate that exercise-related adaptations can differ across outcomes. However, differences in programme content, participant characteristics, and measurement methods limit direct comparisons with the findings of the current study.

The findings extend the local rationale established by earlier Ugandan research. Nsamba and colleagues identified a higher fat mass index among children and adolescents with T1DM than among matched participants without diabetes (5). The present study moves from describing those differences to examining outcomes following a programme designed to address nutrition, activity, and self-management together. Nevertheless, lower fat mass or BMI should not automatically be interpreted as better health across a population spanning adolescence and young adulthood. Their significance depends on initial nutritional status, growth, and concurrent changes in fat-free tissue.

Several pathways could plausibly contribute to the observed differences. Personalised dietary counselling might improve food selection and meal planning, while supported physical activity might influence energy expenditure and muscle use. Practical demonstrations and continued engagement with diabetes nurses and a nutritionist could also support more consistent self-management. The intervention also combined several components, preventing attribution of any association to an individual element. This distinction matters because behavioural and clinical responses need not occur together. For example, Nansel and colleagues’ family-based nutrition trial improved dietary quality without improving glycaemic control (9).

The circumstances in which participants manage diabetes are also important. Ndahura and colleagues documented substantial gaps in caregivers’ diabetes-related nutrition knowledge, supporting the importance of accessible, practical education (6). However, knowledge is only one influence on self-management. Sseguya and colleagues showed that disruptions to income, food access, and diabetes services affected management practices among young adults in rural Uganda (7). These findings suggest that the effectiveness of education depends partly on whether participants can act on the advice they receive. Adapting counselling to locally available foods and feasible activities is therefore a reasonable design choice, but cultural adaptation alone cannot resolve material constraints.

The principal design limitation is the relationship between clinic attendance and allocation. Participants entered the groups through an alternating clinic-week schedule. Although maintaining separate clinic sessions was intended to reduce contamination, it also linked intervention exposure to clinic-day grouping. Differences in attendance patterns, care delivery, or participant circumstances could consequently contribute to the observed associations. The study also did not account for factors such as pubertal development, diabetes duration, insulin regimen, glycaemic control, or food security.

The study nevertheless has useful features. It evaluated a locally adapted programme within an existing diabetes service, included a concurrent comparison group, and examined several body composition outcomes alongside conventional anthropometry. Assessment at six months extended beyond the eight-week intervention period. The next step is a larger evaluation that can distinguish intervention effects from clinic-level influences and measurement variation. Such a study should use repeated measurement of dietary intake, physical activity, insulin use, glycaemic outcomes, and hypoglycaemia to help establish mechanisms and clinical relevance. Measurements immediately after the programme and at later follow-up would distinguish initial response from maintenance. Functional outcomes, such as strength or fitness, would also help determine whether changes in estimated muscle mass correspond to benefits that matter to participants. Overall, participation in the programme was associated with differences in body composition, including modest changes among intervention participants and declines in fat-free tissue estimates among controls. These findings justify further evaluation of integrated nutrition and physical activity support within Ugandan diabetes services.

## Conclusion

A culturally tailored health promotion programme was associated with lower fat mass and BMI and higher fat-free mass and muscle mass at follow-up among adolescents and young adults living with T1DM in Wakiso, Uganda. Differences reflected modest changes in the intervention group alongside less favourable changes among controls. These preliminary findings support further evaluation of integrated nutrition and physical activity support within diabetes services. The programme’s causal effects, clinical benefits, and suitability for wider implementation remain to be established.

## Funding

This study was funded by a small research grant to Jonathan Nsamba by the Ramani Fund [https://www.ramanifund.org/]. The funder had no role in the study design, data collection, analysis, interpretation, or writing of the manuscript. All authors take responsibility for the integrity of the work and the decision to submit for publication.

## Acknowledgment

We thank the adolescents and young adults who participated in this study and the caregivers who supported their participation. We are grateful to the staff of Wakiso Health Centre IV for facilitating recruitment and data collection. We also acknowledge the research team members who supported participant assessments and data management, the Changing Diabetes in Children programme, and Hope Springs Health Foundation Uganda for its institutional and logistical support.

## Author Contributions

Conceptualisation: JN, WS, SG

Data Collection: JN, RB, KHA

Data Analysis: JN, NM, KHA

Original Draft: JN

Review & Editing: SG, CM, SA, RT, WS, NM, KHAN

Supervision: WS

Funding Acquisition: JN, SG

All authors reviewed and approved the final manuscript and agree to be accountable for the integrity of the work.

## Data Availability

The data are not publicly available because of confidentiality restrictions. Requests for access to de-identified data may be directed to the corresponding author and will be considered subject to applicable ethical approvals and data-protection requirements.

## What is already known on the topic

1. Previous Ugandan research has identified differences in body composition between young people with T1DM and participants without diabetes, alongside gaps in caregivers’ nutrition knowledge.
2. Nutrition and physical activity interventions can improve selected behavioural and health outcomes, but findings from other settings do not establish their effects within routine Ugandan diabetes services.

## What this study adds

1. A culturally tailored programme delivered within an existing Ugandan diabetes clinic was associated with differences in estimated body composition at six months.
2. The differences reflected modest changes among intervention participants and declines in fat-free tissue estimates among controls.
3. The findings provide preliminary evidence for a larger randomised evaluation.

## Notes

### Competing Interest Statement

The authors have declared no competing interest.

### Author Declarations

Ethical approval was obtained from the St. Francis Hospital Nsambya Research Ethics Committee (SFHN-2024-133).

